# Understanding digital health use among postpartum women in Alberta, Canada: a qualitative focus group study

**DOI:** 10.64898/2026.04.26.26351785

**Authors:** Viktoriia Kurkova, Setayesh Modanloo, Yiqing Wu, Julie Tian, Emilie Desnoyers, Medard K. Adu, Gina Wong, Andrew Greenshaw, Jake Hayward

**Author notes:** Corresponding Author:, (VK).

## Abstract

The postpartum period involves profound physical, emotional, and social changes, yet many women report fragmented, infant-centered care that leaves their own needs insufficiently addressed. Digital health tools, including mobile apps, wearables, telehealth, and online resources, are increasingly used by postpartum women to seek information, support, and reassurance; however, little is known about how women experience these tools in their everyday lives. This qualitative study employed thematic analysis to explore the perspectives of postpartum women on digital health. Postpartum women (≤12 months after birth) living in Alberta, Canada, were recruited through maternity clinics and targeted social media advertisements. Four virtual focus groups (4-6 participants in each; 18 participants overall) were conducted via Zoom using a semi-structured guide on postpartum healthcare experiences, use of digital tools (apps, wearables, telehealth, AI), and perceived barriers and facilitators to adoption. Sessions were audio-recorded, transcribed verbatim, and coded by multiple researchers.

Thematic analysis identified 32 codes, organized into 12 subthemes and four overarching themes: navigating postpartum support networks; empowerment through digital health tools; conditions for acceptable digital health design; and when technology feels like a burden. Women appreciated multiple sources of support from midwives, public health nurses, peers, and online communities, but described care that quickly became infant-focused, leaving their own recovery and mental health under-addressed, particularly in rural settings. Digital tools helped mothers structure infant and self-care, track symptoms, and prepare for appointments, yet also created new forms of burden, including information overload, usability challenges, privacy concerns, and feelings of surveillance or pressure to perform. Participants emphasized personalization (flexible notifications, mother-focused content), embedded mental health support, integration with trusted providers, and co-designed, credible platforms endorsed by Canadian health systems. Overall, to be acceptable and effective, tools must center mothers’ needs and be embedded within a broader ecosystem of responsive, continuous care.

**Author summary:** Becoming a parent is a major life change, and many women feel that support from the health system drops off once the baby is born. At the same time, new mothers are increasingly turning to mobile phone apps, wearable devices, online groups, and video visits to answer questions, track health, and feel less alone. We wanted to understand women lived experience with these digital tools after giving birth: what feels helpful, what feels burdensome, and what they would want in an ideal tool.

Our research team, consisting of three PhD students, held four online focus group discussions (4-6 participants per group; 18 participants overall) with women in Alberta, Canada, who had given birth within the past year. They described digital tools as both empowering and exhausting. Apps and wearables helped them track feeding, sleep, and symptoms, organize daily life, and come better prepared for medical appointments. At the same time, constant tracking, frequent notifications, and unclear data practices could feel overwhelming, guilt-inducing, or intrusive.

This study is an important first step in a larger co-design work. By listening closely to mothers’ stories, we gathered practical ideas about what a supportive postpartum app should (and should not) do. In future phases, we plan to work directly with postpartum women and frontline clinicians to turn these ideas into a user-friendly, trustworthy digital tool that supports both mothers’ and babies’ health.

## Introduction

The postpartum period is a time of significant physical, emotional, and social adjustment after giving birth (1). In clinical and public health literature, it is often defined as the first 6 weeks after birth, although recovery and adaptation frequently extend beyond this window (2). During this time, new mothers undergo complex transitions as they adapt to the demands of parenthood (3). This critical phase is marked by heightened vulnerability; many women experience significant emotional strain, including symptoms of anxiety and depression, while simultaneously developing essential parenting skills (4).

In Canada, 23% of recent mothers report symptoms consistent with postpartum depression or anxiety disorder (5). However, despite the clinical and psychosocial significance of this period, there are well-documented gaps in postpartum care. A 2021 survey of 561 Canadian mothers showed that 20% were unsatisfied with their postnatal care, and 3.2% had no postnatal visits (6). Because an estimated one in five Canadians lacks a regular primary care provider, emergency departments (ED) often become the only accessible option for postpartum women (7). A retrospective cohort study from Alberta found that nearly 45% of women visited the ED within the first year after delivery, yet only 5.2% of these visits resulted in admission, suggesting many encounters may have been avoidable or not medically necessary (8). Rural residence, younger maternal age, and pre-existing mental or physical health conditions were associated with higher ED use. Taken together, these patterns highlight gaps in the postpartum care model, where timely, appropriate support is often lacking, and some ED visits might be preventable if reliable information and guidance were more readily available.

The growing integration of digital health solutions presents compelling opportunities to bridge these gaps. Telehealth and remote monitoring can reduce logistical barriers for women who are home with newborns, juggling caregiving demands, or living far from services. For example, in a retrospective cohort study, implementation of virtual postpartum visits was associated with higher attendance at follow-up appointments and increased rates of postpartum depression screening (9). Similarly, a text-based remote blood pressure monitoring program for individuals with hypertensive disorders of pregnancy led to lower postpartum readmission rates, fewer emergency department visits, and reduced overall medical costs within six months of delivery (10).

While these findings illustrate the potential of digital modalities to improve accessibility and safety, gaps persist in understanding how these technologies are experienced, navigated, and adopted by women. Prior work has emphasized the need for deeper, experience-based insights into the types of content new mothers find most valuable, the ways they engage with digital platforms, and the barriers that prevent sustained use (11). Without this contextual understanding, many digital health interventions risk failing to meet the real-world needs of the populations they intend to serve (12). This study aims to address this gap by exploring digital health use among postpartum women in Alberta, focusing on their experiences with healthcare access and their perspectives on digital tools for self-monitoring and support.

## Methods

### Study design

This qualitative focus group study was informed by Braun and Clarke’s thematic analysis framework, emphasizing flexibility, reflexivity, and researcher transparency throughout the analytic process (13).

### Study setting

The study was conducted in Alberta, Canada. All data collection took place online, with focus groups held virtually to enable participation from both urban and rural regions of the province.

### Participants and sampling

Participants were postpartum women residing in Alberta within one year of giving birth. We included women up to 12 months postpartum because physical recovery, service navigation, and mental health experiences (including depressive and anxious symptoms) can persist or emerge across the first postpartum year, and later-postpartum participants can reflect on what would have been most helpful earlier while still experiencing ongoing needs. We used purposive sampling to include women with varying levels of digital health use and living in both urban and rural settings. Rural residence was defined based on geographical proximity to urban services, operationalized as living one hour or more travel time from a major urban centre.

### Recruitment procedures

Recruitment was conducted over a two-month period through maternity clinics and targeted social media advertisements (Instagram and Facebook). Recruitment materials briefly described the study aims, eligibility criteria, and what participation would involve, and directed interested women to contact the study team for screening and consent. In total, 19 women were screened and enrolled; one participant did not attend any focus group session and did not respond to follow-up messages, resulting in a final sample of 18 participants. All participants received a 15 CAD Walmart gift card as a token of appreciation for their time.

### Data collection

Using focus groups as a qualitative method, we sought to capture women lived experiences, perceived gaps in existing postpartum support, and preferences in digital health. A semi-structured discussion guide (Supplementary file 1) was developed, drawing on an informal review of the literature on attitudes toward digital health and consultations with researchers specializing in postpartum health. The guide covered five broad topic areas: (1) experiences with accessing postpartum healthcare and support; (2) prior and current use of digital health tools; (3) perceived barriers to using these tools; (4) factors that might make digital health tools more acceptable or helpful; and (5) perceived impact of digital health on women’s overall healthcare experience. Before data collection, the guide was pilot-tested with members of the research team acting as participants, and minor adjustments were made to question wording and ordering based on this feedback.

Four virtual focus groups were conducted via Zoom, each including four to six participants and lasting approximately 70 minutes. Sessions were facilitated by two researchers (V.K. and S.M.) with support from a co-moderator (Y.W.), following the semi-structured guide. All sessions were audio-recorded and transcribed verbatim, and identifying details were removed from transcripts to maintain participant confidentiality.

### Data analysis

Two researchers (V.K. and S.M.) independently reviewed and coded the first transcript line by line, identifying initial codes inductively to allow codes to emerge from the data. The research team then met to compare interpretations and develop a coding framework, which was used to guide coding of subsequent transcripts and iteratively refined as new codes and nuances emerged, in line with reflexive thematic analysis principles (13). Analysis proceeded until thematic saturation was reached, that is, no substantively new codes or themes were identified in later focus groups, suggesting that the data were sufficiently rich to address the research aims (14,15). Through iterative team meetings, the final coding framework was organized into sub-themes and overarching themes that captured patterns across the dataset. A third researcher (Y.W.) independently reviewed the coded dataset and contributed to resolving discrepancies and consolidating themes, enhancing analytic rigor through triangulation and team discussion (16). V.K. and Y.W. are clinicians whose practice experience informed their sensitivity to the clinical realities of postpartum care, while S.M. is a psychologist with formal training in thematic analysis whose expertise supported methodological rigor during coding and theme development.

### Ethical approval

The study protocol was reviewed and approved by the University of Alberta Health Research Ethics Board (Pro00147501). All participants provided informed consent prior to taking part in the focus groups, and all procedures followed relevant institutional and national ethical guidelines for research involving human participants.

## Results

A total of 18 women took part in the study. All participants identified as cisgender and ranged in age from 22 to 41 years (median = 32). Participants were recruited from both urban (n = 13) and rural (n = 5) regions of Alberta, offering a wide range of perspectives regarding healthcare access and digital literacy. The focus group discussions included mothers who regularly used mobile health applications and wearable devices (n = 16), as well as those with limited exposure to such technologies (n=2).

### Overview of Themes

Thematic analysis identified a total of 32 thematic codes, which were further organized into 12 sub-themes. These sub-themes were then grouped into 4 overarching themes (Fig 1). A visual overview of the themes and sub-themes is presented in Fig 1, and a detailed interpretation of each sub-theme, including core meaning and illustrative focus, is provided in Table 1.

**Fig 1.**
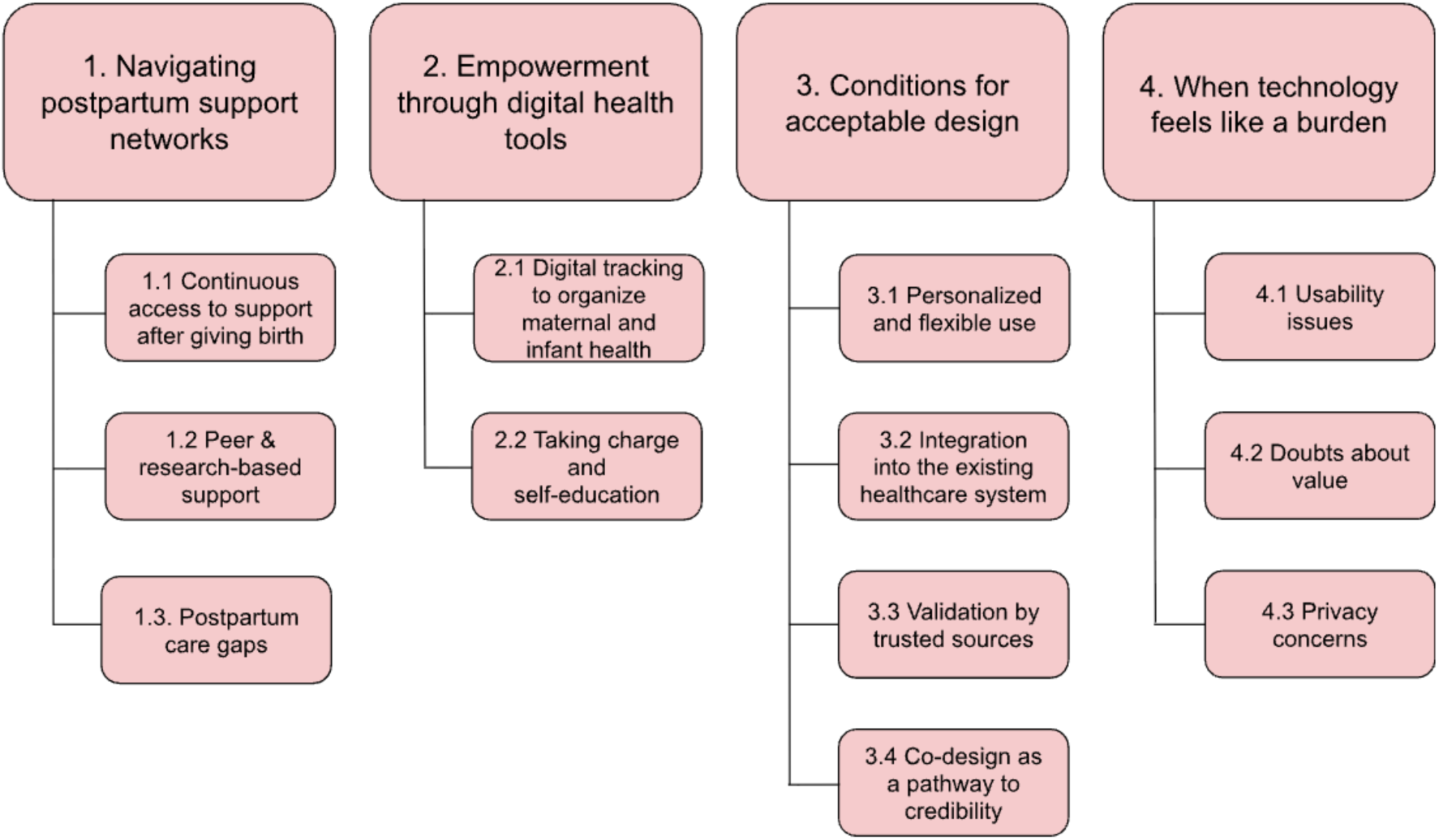
Overview of themes.

**Table 1:** Interpretation and illustrative focus of sub-themes.

| Theme | Subtheme | Core meaning/interpretation | Illustrative focus |
| --- | --- | --- | --- |
| Navigating postpartum support networks | Continuous access to virtual support after birth | Ongoing hybrid (virtual + in-person) contact with midwives, nurses, and portals created reassurance and reduced uncertainty between visits | Telehealth, messaging, hotlines, home visits |
|  | Peer & research-based support | Online communities and structured programs provided emotional validation, normalization, and stage-specific guidance | Facebook/WhatsApp groups; text-based research supports |
|  | Postpartum care gaps | Care became infant-centered; mothers felt overlooked and had to self-advocate, especially in rural areas; information overload increased anxiety | Long travel, fragmented care, “Dr. Google,” conflicting advice |
| **Empowerment through digital health tools** | Digital tracking to organize maternal and infant health | Apps and wearables functioned as memory aids, reducing cognitive load and enabling shared caregiving and preparation for appointments | Feeding/sleep logs, wearables, baby monitors |
|  | Taking charge and self-education | Women used portals and AI tools to interpret information, understand results, and participate more actively in decisions | Summarizing research, accessing lab results, preparing questions |
| **Conditions for acceptable design** | Personalized and flexible use | Tools must adapt to postpartum realities with customizable reminders and equal focus on maternal health and mental health | Notification control, mood check-ins, mother-focused features |
|  | Integration into the existing healthcare systems | Digital tools should connect directly with providers and records to support continuity and reduce visit burden | Secure messaging, data sharing, digital triage |
|  | Validation by trusted sources | Institutional endorsement and evidence-based content increased credibility and willingness to use tools | Provider/government backing |
|  | Co-design for credibility | Involvement of mothers and frontline clinicians enhances relevance, empathy, and trust | Participatory development |
| **When technology feels like a burden** | Usability challenges | Cognitive effort, device discomfort, battery demands, and excessive tracking created fatigue and stress | Learning new tools, constant charging, over-tracking |
|  | Doubts about value | High costs and fragmented apps reduced perceived benefit | Multiple subscriptions |
|  | Privacy concerns | Uncertainty about data storage and infant information sharing limited trust and adoption | Data governance, consent worries |

### Theme 1: Navigating postpartum support networks

#### Subtheme 1.1: Continuous access to support after giving birth

Participants shared their experience transitioning from pregnancy to postpartum care. They valued having reliable, ongoing access to guidance immediately after discharge and overall, in the postpartum period. Mothers (n=12) described using a range of digital supports, including virtual appointments, public-health hotlines, and messaging with healthcare providers through patient portals, alongside in-person nurse visits and midwife follow-ups, especially in the first weeks postpartum. They described feeling more secure and confident when they had reliable, ongoing access to trusted support.

> “One of the great things that I also got was access to my therapist that I was seeing during pregnancy, she was able to see me like virtually and she was totally fine with my baby being fussy during the meeting and all that stuff.” (Focus Group 2, Participant 3)
>
> “I can just send a quick question to my doctor through the patient portal, and I’ll get a response within a day or two. And I have found that helpful for the things that really actually matter to be able to just reach out and not have to rely on Dr. Google. It is immensely helpful. It is immeasurably valuable.” (Focus Group 2, Participant 5)

Midwives’ check-ins and public health nurse outreach were repeatedly (n=4) described as very helpful, which made women feel seen and safe between formal appointments.

> “I really appreciated the public health nurse and home visit to know I was on the right track, and not having to travel with a baby. Any type of virtual visits, especially in the first couple months there, was… just amazing.” (Focus Group 3, Participant 3)

#### Subtheme 1.2: Peer & research-based support

Alongside professional contact, participants highlighted online peer groups (n=5; web-cites forums, WhatsApp groups, Facebook groups) and research studies with structured support provided often through text messages (n=4; SmartMom, P3 cohort) as vital sources of emotional support, reassurance, and a sense of connection during pregnancy and after birth. Some moms preferred to read rather than post, finding validation by seeing others’ experiences.

> “I was part of an Edmonton mothers’ group, and it was a great place to see what others were going through; sometimes just reading others’ comments helped.” (Focus Group 2, Participant 4)

Those who took part in a pregnancy research studies valued stage-specific prompts that arrived at the right time, check-ins, and mental health assessments.

> “I joined the P3 cohort; they screened for postpartum depression and did weekly check-ins based on where you were in pregnancy, and they would give you a resource that you may want to look into during this period.” (Focus Group 2, Participant 10)

#### Subtheme 1.3: Postpartum care gaps

Some women (n=3) felt that once the baby arrived, healthcare services often shifted focus entirely to the infant, leaving them feeling overlooked and without guidance for their recovery. One mother never received her six-week follow-up because the appointment was scheduled on Christmas Eve. This experience was especially challenging for first-time moms. In this context, some mothers (n=3) described having to actively advocate for themselves, including searching online and preparing their symptoms before appointments to ensure their concerns were taken seriously and the right issues were addressed. Moreover, those in rural areas (n=3) faced added barriers, including long travel times and limited services.

> “We live rurally, so I’m driving over an hour each way three times a week for her care. As a first-time mom, I’ve had no time to deal with my own health; the focus is so much on her, and the idea of adding another long trip just for me feels impossible. After the baby was out, it was like I no longer mattered.” (Focus Group 2, Participant 2)
>
> “I like the Alberta app because I can see all my blood test results. Sometimes, if I don’t bring up the right symptom with my doctor or public health nurse, they don’t ask, and we go down the wrong path. I feel like I have to google things first so I know what to tell them before the appointment.” (Focus Group 3, Participant 3)

Participants (n=11) also struggled with an overwhelming amount of inconsistent or conflicting information from various sources, which heightened their anxiety and made it hard to know what advice to follow.

> “It’s so hard to parse through the sea of information. You can find both positive and negative confirmation bias for literally everything.” (Focus Group 2, Participant 5)

### Theme 2: Empowerment through digital health tools

#### Subtheme 2.1: Digital tracking to organize maternal and infant health

Mothers described using a mix of apps, baby devices, and wearables to keep track of both their own health and their baby’s routines. Baby-tracking and milestone apps such as Huckleberry, Baby Tracker, BabyTime, the CDC milestone app, The Bump, What to Expect, Baby Center, BabyChat, and Solid Starts, as well as connected devices like the SNOO smart bassinet, smart socks, and Nanit baby monitors, were used to log feeds, sleep, diapers, medications, and developmental milestones (n=10). For their own health, women relied on wearables and apps, including Fitbit, Apple Watch, MyFitnessPal, and other activity and sleep trackers like Bellabeat Ivy to monitor sleep, steps, heart rate, and nutrition (n=10). Having this information recorded and easily accessible helped them plan their day, share caregiving tasks with partners, and answer questions from health-care providers without relying on memory while sleep-deprived.

For many (n=14), digital tracking reduced mental load and anxiety by making patterns visible and providing concrete, up-to-date information about how they and their babies were doing:

> “I track how much milk I’m producing, and my smartwatch helps me monitor sleep and steps going back to the gym, which is really useful when my memory is garbage from lack of sleep.” (Focus Group 2, Participant 2)
>
> “I’m mostly using smartwatches to track baby things, and I found that really useful to just like have that off my mind. I don’t have to think about it. I just can check when it was the last time he changed or when he sleeping. To be in a piece of mind, I really enjoy using that.” (Focus Group 4, Participant 4)
>
> “My husband can put everything in the app. When I get back after a workout, I know exactly where she’s been in her schedule. I can just kind of seamlessly jump back into caring for her.” (Focus Group 2, Participant 4)

#### Subtheme 2.2: Taking charge and self-education

Mothers described using digital tools not only to receive information, but to actively interpret it and take a more informed role in their care. Some (n=6) turned to AI tools such as ChatGPT or Gemini to summarize research articles and provincial health websites into simpler language they could understand and apply in everyday decisions. Others (n=4) emphasized how immediate access to lab results and visit summaries through platforms like Connect Care or MyHealth Alberta (provincial digital clinical information systems) allowed them to stay informed between appointments, prepare questions in advance, and feel less dependent on clinicians to interpret every detail. Together, these practices reflected a form of digital self-education, where women used technology to “put their health care in their own hands” in a system where they often felt they had to speak up and come prepared to get their concerns addressed.

> “I go through a lot of articles and ask Gemini to summarize or explain them in simpler terms so I can actually understand what the research is saying. I also use it to re-explain information from Alberta Health websites, because it’s written in a way that is not always easy to understand.” (Focus Group 3, Participant 2)
>
> “I love getting my results on my Connect Care. Whenever I have a blood test or any sort of medical test, I get them right away. So I feel like it helps me put my healthcare in my own hands.” (Focus Group 1, Participant 4)

### Theme 3: Conditions for acceptable design

During the focus groups, we asked participants to imagine an “ideal” postpartum digital tool and to describe what would make it genuinely useful for them. Women described a set of conditions for acceptability: tools should be personalized and flexible in everyday use, integrated with existing healthcare services, clearly validated by trusted sources, and developed through co-design with postpartum mothers and frontline clinicians.

#### Subtheme 3.1: Personalized and flexible use

Participants (n=10) consistently expressed the need for personalization in digital health tools like mHealth apps and wearables, so that the technology fits naturally into the unpredictability of postpartum life. They wanted freedom to decide which notifications and reminders they received, when, and how frequently.

> “I think reminders would depend on what exactly I was being reminded about. If it was something like developmental milestones your baby I want it once week. If it’s something as important as take your medication I want that daily.” (Focus Group 1, Participant 2)

Mothers (n=6) highlighted that many existing postpartum mHealth apps are heavily oriented toward the baby, with far fewer options to monitor their own health. Several participants described realizing late in the day that they had not eaten, rested, or checked in with themselves, despite meticulously logging feeds, diapers, and sleep for their infant. As a result, they expressed a clear preference for tools that give equal attention to maternal and infant health.

> “There’s so much to track for your baby, but then so little to track for yourself. Apps all seem very focused on baby, not mama. So I think something that would help moms specifically would be really important.” (Focus Group 2, Participant 5)

Women (n=8) also wanted mental health supports built into postpartum apps, including simple mood check-ins, tailored information about postpartum emotional changes, and quick routes to appropriate services or self-help strategies.

> “I really want an app to be able to do something with the emotions of a postpartum person. Having like a shortened five-question questionnaire that’s just you know checking in on me would be really nice.” (Focus Group 2, Participant 6)
>
> “I definitely think there should be an ideal app, something where I can go and check, like, hey, I feel this way. And they have some exercises or some reasoning why you feel that way” (Focus Group 4, Participant 2)

Another preference (n=5) was inclusivity for family members and partners. Mothers emphasized that their postpartum journey involves a network of support, and digital tools should reflect this. They wanted the ability to share specific information, such as feeding logs or reminders, with trusted caregivers while maintaining control over what remains private.

> “Information sharing to your partner being accessible, or even customize a list of people beyond your partner if you have a support group, like a sister, a mother, an aunt, would be really helpful.” (Focus Group 2, Participant 5)

#### Subtheme 3.2: Integration into the existing healthcare system

Mothers described how it can be difficult to absorb complex advice from their healthcare provider when they are tired, emotional, or overwhelmed during postpartum appointments. Mothers wanted the app to act as a bridge to their care team. They wanted a tool that would allow them to access visit-related information at their own pace and revisit it whenever needed, rather than relying on memory or scattered notes. Half of the participants (n=9) envisioned an integrated portal or secure messaging feature that would connect directly to their family physician, midwife, or another provider. Several participants (n=5) also noted that being able to share their tracked data (for example, symptoms or baby routines) ahead of appointments would help them remember what to raise and give providers a clearer picture of what had been happening at home.

> “I want an app that is integrated with our healthcare system. This would be so much easier if the doctor could just access what I’m tracking for my baby. I would give permission.” (Focus Group 4, Participant 3)
>
> “I like being able to review the information at my leisure, not like all this information overload when you’re at your healthcare provider, who has a limited amount of time.” (Focus Group 1, Participant 3)

Finally, participants (n=4) expressed a strong interest in digital triage or pre-visit assessment to minimize time in waiting rooms and reduce infection exposure for vulnerable infants.

> “Being able to triage at home, and then for them to send you a text message to get you in and out faster than just sitting in that cesspool of a waiting room.” (Focus Group 3, Participant 1)

#### Subtheme 3.3: Validation by trusted sources

Trust was a deciding factor in whether participants would use any type of digital health app. Mothers expressed greater confidence in tools endorsed by healthcare providers (n = 4) or developed under recognized Canadian or governmental bodies (n = 4). Professional validation signaled safety and reliability.

> “I’d want to know that the information in the app is from a really reputable source; if my doctor vouches for it, I trust it.” (Focus Group 1, Participant 3)
>
> “I trust Canadian and/or government ones more than private; I don’t believe they’re going to sell your data.” (Focus Group 3, Participant 1)

#### Subtheme 3.4: Co-design as a pathway to credibility

Participants also emphasized that trust in digital health apps depends on who is involved in creating them. They advocated for the involvement of both postpartum mothers (n = 3) and frontline clinicians (n = 4), particularly nurses and doctors, in developing and reviewing content to ensure that it is practical, empathetic, and clearly communicated.

> “It is important to engage moms who’ve gone through it to see what’s really useful to them.” (Focus Group 2, Participant 6)
>
> “Maybe it’s nurses that need to be the in-between—to bridge that gap and make sure people interpret the information correctly.” (Focus Group 3, Participant 1)

### Theme 4: When technology feels like a burden

While self-tracking provided structure and reassurance for some, others described it as emotionally and physically exhausting. Participants discussed three aspects of burden: usability issues, doubts about value, and privacy concerns.

#### Subtheme 4.1: Usability issues

Adopting new technology was described as cognitively taxing, particularly when participants were sleep-deprived or overwhelmed with infant care (n = 3).

> “Adopting new technology is hard—there’s just too much to figure out when you’re already tired and taking care of a baby.” (Focus Group 2, Participant 3)

Short battery life reduced motivation to engage for some participants (n = 3).

> “Battery life is a huge thing; I have to charge my watch almost twice a day. That’s really annoying.” (Focus Group 1, Participant 4)

Several women (n = 2) reported that wearables were physically uncomfortable or interfered with caring for their baby.

> “I would stop wearing my Fitbit, because I don’t want this digging into baby’s back, I don’t want it to be uncomfortable.” (Focus Group 4, Participant 2)

Some mothers (n = 10) felt pressure to track every detail of their baby’s care, such as feeding, sleep, and diapers, in order to feel in control or to meet expectations of “doing things right.” This constant tracking often became overwhelming and could take away from simply being present with their baby:

> “I feel like I have to track everything now and can’t just go with the flow. If I don’t track it, it’s like it doesn’t count.” (Focus Group 2, Participant 5)

In addition, many devices and apps weren’t tailored to their unique needs at the postpartum stage, for example, giving discouraging messages about weight gain or decreased physical activity that are actually normal during recovery:

> “So even just updating my weight to postpartum, the app was like ‘you’ve gained a lot of weight,’ and then nagging me about not getting enough steps. I just had a baby. I’m still recovering. Thank you for making me feel horrible about that. There’s nowhere to track that or to say what’s normal at six weeks postpartum.”(Focus Group 3, Participant 4)

#### Subtheme 4.2: Doubts about value

Mothers (n = 6) were frustrated by high subscription costs and the need to use multiple apps for different purposes.

> “A lot of the apps require a subscription, and then not knowing if it’s actually worth your money or not. And the really expensive often, like, for a year subscription, when you’re on maternity leave, like, none of us can afford that, so… yeah, that’s frustrating.” (Focus Group 3, Participant 4)
>
> “I don’t really want to spend $200 a year on ten different apps; none of them do everything I need.” (Focus Group 3, Participant 4)

#### Subtheme 4.3: Privacy concerns

Some moms (n = 3) weren’t worried about privacy when using apps or devices, saying it’s become normal for phones and technology overall to collect data. They saw it as something inevitable in today’s world.

> “If you use any sort of digital tool, your information is out there. They’re not going to be using *any more information than what is already out there on the web.” (Focus Group 4, Participant 1)*

Other mothers (n = 13) expressed concerns about how their personal or health data might be accessed, stored, or shared by apps and digital platforms, especially if they are not Canada-based. They questioned who would see the information and whether it was truly secure, leading to hesitation about what they were comfortable sharing. Some felt uneasy about vague consent language and even decided not to use the tool after reading the terms and conditions.

> “I’m 100% worried about how my or my baby’s health data might be used, that’s why I don’t use apps!” (Focus Group 3, Participant 4)
>
> “I bought a baby monitor that could connect to my phone, but after reading the app’s privacy terms, I decided not to put it on WIFI because the data use sounded too broad and open-ended.” (Focus Group 2, Participant 5)

## Discussion

This study offers an in-depth account of how postpartum women navigate digital health tools within a broader landscape of fragmented, infant-centered care. Women described apps, wearables, online communities, and virtual visits as extending support between clinical encounters, helping them organize infant and self-care and feel less alone, while also introducing new forms of burden and worry. Rather than seeing women as simply eager or resistant users, our findings show that they weigh potential benefits against effort, emotional impact, and trust. This suggests that the promise of postpartum digital health depends on how well tools are designed for and embedded in women’s everyday realities after birth.

Across health systems, postpartum care often shifts quickly from a focus on the pregnant person to a focus on the infant, with persistent gaps in maternal follow-up. Mothers describe feeling left out and having unmet information needs about their own recovery and mental health, reinforcing the sense that the system’s attention moves away from them once the baby is born (17,18). In response, many women assemble “hybrid” support networks that combine in-person and virtual contact with healthcare providers, alongside structured peer programs and informal online communities. Qualitative studies show that mothers value opportunities to connect with both professionals and peers across the first postpartum year, and that peer forums and group supports can be critical spaces for validation, stigma reduction, and sharing practical strategies, particularly for those experiencing postpartum depression or distress(19–21). These findings suggest that digital tools can act as conduits that connect mothers to multiple forms of support when formal care is time-limited or difficult to access.

In our focus groups, digital tools played an organizational and interpretive role in women’s everyday routines. Apps, smart monitors, wearables, and portals helped track infant feeding and sleep, as well as maternal sleep, activity, and symptoms, giving concrete records that could be used to answer questions, notice patterns, and feel more prepared for appointments. Evaluations of postnatal apps and infant-feeding trackers report similar benefits, with mothers describing tracking and stage-specific information as reassuring during the transition to parenthood and helpful for structuring conversations with providers (22,23). Perinatal digital health studies likewise describe women using apps, portals, and web resources to understand test results, track symptoms, and feel more in control of their and their child’s care (11,24). This sense of control can strengthen parental self-efficacy, understood as a parent’s belief in their ability to manage parenting tasks and positively influence their child’s development, and higher parenting self-efficacy is consistently linked to fewer depressive symptoms(25). In the mental health space, ecological momentary assessment and mood-tracking apps have been perceived by perinatal women and providers as promising for detecting emerging difficulties between visits, particularly when feedback is easy to interpret and connected to clear follow-up options (26). Together, this suggest that postpartum women already rely on digital tools to lighten the mental load of managing care and to advocate for themselves within time-limited, often fragmented services.

At the same time, the content and configuration of current tools often did not match what women felt they needed. Similar to our findings, analyses of postnatal apps found that fetal and infant development content is common, while information on maternal self-care and psychological well-being is comparatively sparse or fragmented (27). Women in our study described wanting brief mood check-ins, simple explanations of what emotional changes are typical versus concerning, and clear paths to support, which aligns with evidence that low-burden mood monitoring and psychoeducation can increase awareness and prompt earlier help-seeking when embedded in appropriate care pathways (26,28,29). Personalization and flexibility were also key: women wanted to decide what to track, how often reminders appear, and what could be shared with partners or other caregivers. Evaluations of infant-feeding and parenting apps show that context-specific prompts, customizable features, and clear evidence-based content are associated with perceived relevance and sustained engagement (30,31).

Additionally, preferred future tools in our focus groups were integrated with existing health portals, endorsed by Canadian or governmental bodies, and co-designed with postpartum women and frontline clinicians, which is consistent with maternal mHealth research showing that participatory, evidence-informed, and institutionally backed apps are perceived as more credible and usable(32,33). Co-design with mothers, researchers, and healthcare teams is therefore not only a technical approach to improving usability, but also a way to ensure that postpartum tools reflect mothers’ priorities while remaining clinically useful for providers. It can also make data and design choices more transparent, which is particularly important in light of the burdens, pressures, and privacy concerns women described in relation to existing technologies.

Women in our study underscored that digital health tools could intensify, rather than ease, the demands of the postpartum period. Some found it hard to adopt and sustain use of new apps or devices while sleep-deprived and busy with infant care, while others felt pressured to log everything or were discouraged by generic prompts that did not reflect normal postpartum recovery, which sometimes increased guilt and self-criticism instead of reassurance. Studies of postpartum and perinatal self-monitoring tools report similar patterns, with high tracking demands and rigid targets linked to information overload, technostress, and feelings of surveillance or failure (34–36). The postpartum period is a moment of reduced capacity: fragmented sleep, recovery from birth, and a sharp rise in caregiving workload, during which poorly designed digital tools add workload without proportionate benefit, helping to explain the non-adoption and abandonment patterns reported across perinatal mHealth (12).

Privacy and data-governance concerns, particularly around infant information in commercial apps, added a further layer of strain. This echoes work on the “datafied child,” which highlights how routine tracking practices (e.g., sleep, feeding, growth, milestones) can create extensive digital records about children with limited transparency or parental control, raising ethical concerns about surveillance and data use (37). Addressing these practical and ethical burdens is therefore essential if postpartum digital tools are to support, rather than undermine, maternal wellbeing and to remain acceptable and sustainable for women who are already navigating a demanding and vulnerable period.

This study represents an important first step in broader co-design research. The co-design of digital health interventions typically begins with user-centered groundwork, such as in-depth qualitative research to understand users’ needs, priorities, and everyday contexts, before moving into participatory design activities and iterative prototyping(38,39). The focus group data presented here provide a practical foundation for that process by outlining concrete expectations for content, functionality, integration, and data governance from the perspective of postpartum women. Future phases should build on these findings to work directly with postpartum mothers, clinicians, and health-system partners to co-develop, prototype, and evaluate a mother-centered digital health platform that is embedded in routine postpartum care pathways.

### Limitations and future research

This study has limitations that should be considered when interpreting the findings. First, we did not collect sociodemographic variables such as income and race/ethnicity, which constrains our ability to examine how social determinants shape postpartum women’s access to, and experiences with, digital health tools. Preferences, perceived usefulness, and navigation of digital health and support networks can differ for women in various socioeconomic situations. Our sample did not include several groups known to face distinctive postpartum challenges, such as women at high risk for postpartum depression, those with complex medical conditions, Indigenous mothers, or immigrant women with limited English proficiency. In addition, all participants had at least some access to smartphones and internet, so our findings do not capture the perspectives of women who are digitally excluded or have very limited connectivity or device access. Relatedly, a degree of self-selection bias is likely: women who agreed to participate in virtual focus groups are probably more digitally literate and comfortable with technology than the general postpartum population, which may skew findings toward more positive views of digital tools. Although our sample included women within 12 months after giving birth, we did not differentiate between early and late postpartum stages. Needs, stressors, and digital behaviors change substantially between the first six weeks and later months, and these differences may meaningfully shape experiences with digital tools.

Next, albeit our analysis was informed by Braun and Clarke’s reflexive thematic analysis, this methodological approach has inherent limitations. Thematic analysis is an interpretive, researcher-driven method. Hence, our results reflect one of several plausible readings of the transcripts and are shaped by the various theoretical lenses, social positions, and disciplinary backgrounds of the research team (13,40). Within this reflexive approach, we engaged in team-based discussion of how our positions might influence coding and theme development; however, more formal strategies, such as explicitly documenting researcher positionality or maintaining reflexive journals throughout the analysis, could further enhance transparency and provide an additional resource for critically assessing our interpretations (16,41). While we used multiple coders and extensive discussion during our inductive semantic, these strategies do not completely eliminate subjectivity or guarantee that all minority or less vocal perspectives were fully captured, particularly in a focus group environment where group and social dynamics may influence what participants choose to share (16). Future research can include deliberate sampling of underrepresented participants, combining focus groups with one-to-one interviews, or mixed-method designs to strengthen the completeness of the study.

Future research can deliberately sample underrepresented participants, combine focus groups with one-to-one interviews, and use mixed method designs to strengthen the completeness of the study. It will also be important to explore stage-specific patterns of adoption and burden across the first postpartum year, and to strategically recruit more socioeconomically and diverse samples to investigate equity-related differences in digital health adoption among postpartum women.

## Conclusion

The postpartum period in this study was characterized by both meaningful support and notable gaps in care, particularly as attention shifted from mothers to babies and as rural women faced limited local services. In response, participants increasingly relied on digital tools like apps, wearables, online communities, and telehealth to seek information, reassurance, and continuity of care. These tools were experienced as both enabling and burdensome: they supported self-monitoring and self-advocacy, yet also added cognitive load, privacy concerns, and, at times, feelings of self-judgement. Trust in tools was closely tied to credible information sources, transparent data practices, and visible links to Canadian health systems. Taken together, these findings suggest that postpartum digital tools should be developed as part of a broader support ecosystem and co-designed with postpartum women and frontline clinicians. Specifically, clinicians should proactively recommend evidence-based digital tools and discuss data-privacy considerations with postpartum patients as part of routine follow-up, and app developers should prioritize maternal self-care and mental-health features alongside infant-tracking content. The present study offers a user-informed foundation for such co-design work and for future development of equitable, responsive postpartum digital health interventions.

## Data Availability

All relevant data are within the manuscript and its Supporting Information files.

## Acknowledgement

The authors would like to thank Dr. Margie Davenport for her support with participant recruitment. We also sincerely thank all the women who generously shared their time and experiences by participating in this study.

## Author Contributions

Viktoriia Kurkova: Conceptualization, Project Administration, Data Curation, Formal analysis, Writing – original draft; Setayesh Modanloo: Conceptualization, Data Curation, Formal analysis, Writing – original draft; Yiqing Wu: Formal analysis, Writing – original draft; Julie Tian: Validation, Writing – original draft; Emilie Desnoyers: Writing – original draft; Medard Adu: Writing – review & editing; Gina Wong - review & editing; Andrew Greenshaw: Conceptualization, Methodology, Writing – review & editing, Supervision; Jake Hayward: Conceptualization, Methodology, Writing – review & editing, Supervision.

